# Single-Acquisition Spectral and Ultra-High-Resolution Pediatric Head CT: Phantom Comparison of Deep-Silicon Photon-Counting and Energy-Integrating Detectors

**DOI:** 10.64898/2026.10.02.26364533

**Authors:** Samuel G Feemster, Ali Rashidi, Navid Azimi, Zhye Yin, Lusik Cherkezyan, Meghan Yue, Brian E Nett, Amir Pourmorteza

## Abstract

**Background:** Deep-silicon photon-counting CT uses a detector architecture distinct from cadmium-telluride systems and has not been evaluated for pediatric head imaging.

**Objective:** To compare attenuation-dependent performance of deep-silicon photon-counting CT (dSi-PCCT) with energy-integrating detector CT (EID-CT) and assess spectral and ultra-high-resolution (UHR) reconstructions from one acquisition.

**Materials and methods:** Graded-thickness cranial attenuator and pediatric head phantoms were scanned at a matched target volume CT dose index of 18 mGy. dSi-PCCT used 120 kVp and generated 50-, 65-, and 100-keV images; EID-CT used 80 and 100 kVp. We measured CT-number bias, noise, contrast, and contrast-to-noise ratio (CNR) across 0–9 mm of nominal skull thickness. Mixed-effects models assessed imaging-condition effects. We compared matched standard-resolution and UHR temporal-bone images and generated calcium- and water-density maps from the same dSi-PCCT scan.

**Results:** Noise increased by 4.54%, 3.07%, and 1.35% per millimeter at 50, 65, and 100 keV, respectively, versus 8.38% and 7.84% at 80 and 100 kVp. Corresponding CNR decreases were 4.17%, 3.25%, and 1.75% versus 8.94% and 8.54% (interaction *P* < 0.001). Changes in absolute CT-number bias and contrast did not differ among conditions (*P* = 0.92 and *P* = 0.59). In the pediatric head phantom, model-estimated 50-keV CNR ratios were 1.299 (95% CI, 1.163–1.452) versus 80 kVp and 1.424 (95% CI, 1.275–1.592) versus 100 kVp. Across three temporal-bone edges, UHR transition widths were 58.6%–61.9% narrower than standard-resolution widths.

**Conclusion:** dSi-PCCT maintained CNR more consistently as nominal skull thickness increased and provided complementary spectral and UHR pediatric head images from one acquisition.

## INTRODUCTION

Noncontrast head CT is a rapid first-line examination for pediatric trauma, hydrocephalus, shunt evaluation, hemorrhage, and other acute neurologic indications. Brain assessment depends on small gray–white matter differences and stable CT numbers, whereas evaluation of the skull base and temporal bone favors small voxels and sharp reconstruction kernels. These tasks therefore impose competing demands on image noise, contrast, and spatial resolution.

The skull further complicates quantitative brain CT. Beam hardening alters the measured attenuation of intracranial materials, and virtual monoenergetic imaging can reduce this effect ^1^. In photon-counting head CT, keV selection and reconstruction strength affect signal, noise, and contrast-to-noise ratio (CNR), while the calvarium affects measured attenuation in adjacent brain tissue ^2–4^. This is particularly relevant in children because calvarial thickness and head size change with development ^5^.

Photon-counting detectors register individual x-ray events and preserve energy information^6^. Prior dose-matched human brain imaging demonstrated higher gray–white matter CNR with photon-counting than energy-integrating detector CT ^7^. Photon-counting ultra-high-resolution (UHR) imaging has also improved the depiction of small temporal-bone structures ^8–11^. However, published clinical experience with photon-counting CT has predominantly involved cadmium telluride detectors. Deep-silicon photon-counting CT instead uses edge-on silicon detector modules with pixels segmented in depth, a distinct architecture that distributes count rate and supports multibin spectral acquisition ^12,13^. Because this newer detector design may have different imaging characteristics, its performance should be evaluated independently rather than inferred from cadmium telluride systems, particularly for pediatric head imaging.

We compared the attenuation-dependent quantitative performance of deep-silicon photon-counting CT (dSi-PCCT) with energy-integrating detector CT and assessed whether a single dSi-PCCT acquisition could provide complementary spectral and UHR reconstructions in a pediatric head phantom. We hypothesized that dSi-PCCT would maintain CNR more consistently than energy-integrating detector CT as skull thickness increased.

## METHODS

### Study Design

This phantom study compared a 510(k)-cleared, precommercial deep-silicon photon-counting CT system (Photonova Spectra, GE HealthCare, Waukesha, WI) with an energy-integrating detector CT system (Revolution APEX, GE HealthCare). Two linked experiments assessed quantitative performance across controlled attenuation and in an anatomically realistic pediatric head geometry. No living participants were imaged.

### Phantoms

Graded-thickness phantom. A custom cylindrical concrete attenuator surrounded a 1000-mL straight-sided, wide-mouth laboratory jar (Nalgene, Thermo Fisher Scientific, Waltham, Mass; 112-mm outer diameter) containing ten 15-mL Falcon tubes (Figure 1). The attenuator provided a controlled increase in x-ray attenuation intended to model the effect of increasing pediatric skull thickness; its physical thickness was not interpreted as an anatomical skull-thickness measurement. Nominal phantom thickness (hereafter, skull thickness) increased approximately 0.12 mm per 1 mm along the caudocranial axis. Analyses were restricted to 0–9 mm of skull thickness to exclude the terminal phantom and holder regions. The vials contained water, heuristic sodium chloride mixtures, vegetable oil, diluted ethanol, or ferrous sulfate and spanned approximately −120 to +120 Hounsfield units (HU). They were attenuation references rather than chemically calibrated tissue substitutes.

**Figure 1.**
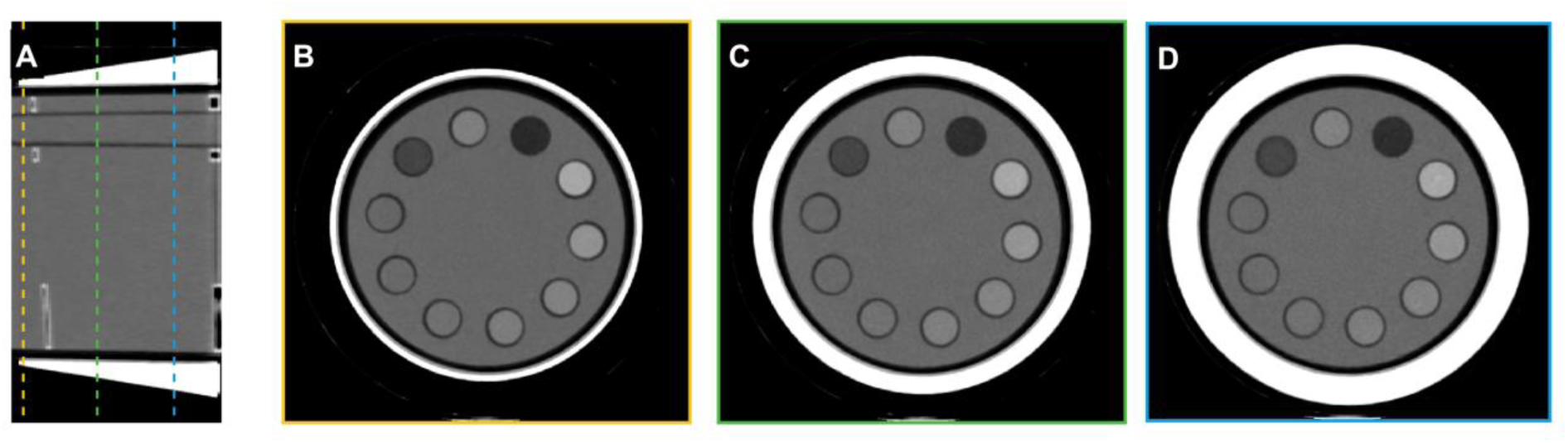
Graded-thickness cranial attenuator phantom. (A) Longitudinal 65-keV dSi-PCCT reformation of the full 72-slice volume. Colored dashed lines mark the three axial positions displayed in B-D. (B-D) Axial slices at nominal skull thicknesses of 1.25, 5.02, and 8.93 mm, respectively. The border colors in B-D correspond to the sampling-position lines in A. Images are displayed at a window level/width of 50/500 HU. Abbreviations: dSi-PCCT, deep-silicon photon-counting CT; HU, Hounsfield unit.

Pediatric head phantom. A commercially obtained cadaveric pediatric skull (supplier-reported age, 4–6 years; Skulls Unlimited International) was filled with ballistic gel and contained eight 2-mL vials from the same general material classes at different dilutions. The phantoms were analyzed independently; only water and fat were directly matched.

### CT Scanning and Reconstruction

Each phantom was scanned with energy-integrating CT at 80 and 100 kVp and dSi-PCCT at 120 kVp. All scans used approximately 40-mm collimation and scanner-reported volume CT dose index near 18 mGy; tube-current–time product was not matched. The dSi-PCCT scan generated complementary 50-, 65-, and 100-keV virtual monoenergetic images. Standard-resolution quantitative images used a 512 × 512 matrix, 170-mm display field of view, and the Standard kernel (MTF50/MTF10, 3.8/6.1 lp/cm). All reconstructions used medium-strength deep-learning reconstruction (DL-M). Slice thickness/increment were 1.25/1.25 mm for the graded-thickness phantom and 2.5/2.5 mm for the pediatric head phantom. The same pediatric dSi-PCCT acquisition also generated a 65-keV UHR series using a 1024 × 1024 matrix, 170-mm display field of view, prototype high resolution kernel for UHR mode (MTF50/MTF10, 15/24 lp/cm), 0.42/0.21-mm slice thickness/increment, and 0.17 × 0.17-mm in-plane voxels. For the matched temporal-bone comparison, the 65-keV standard-resolution (SR) series used the HD Bone kernel (MTF50/MTF10, 5.1/9.3 lp/cm) with 0.42/0.42-mm slice thickness/increment. Manufacturer-supplied MTF50/MTF10 values denote spatial frequencies at 50%/10% of the modulation transfer function as reported by the vendor. We did not activate optional bone-specific or additional beam-hardening correction on either system. Scan and reconstruction parameters are listed in Table 1.

**Table 1.**
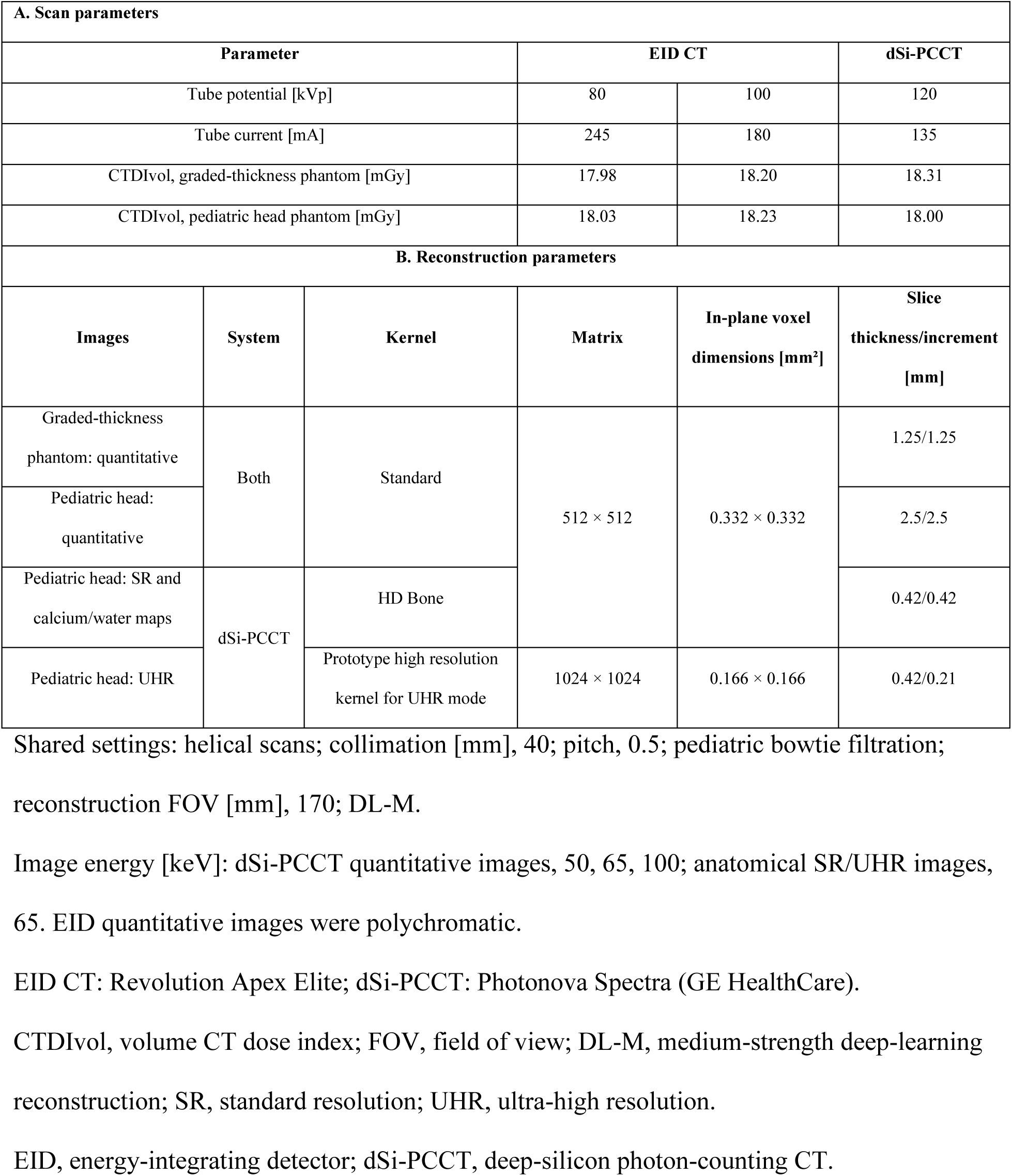
Scan and reconstruction parameters.

### Quantitative Image Analysis

Circular regions of interest (ROIs) were manually placed in each vial with Horos 4.0.0 and propagated through the analyzed slices. Mean ROI area was 32.4 ± 5.0 mm² in the graded-thickness phantom and 20 mm² in the pediatric head phantom. Mean CT number and within-ROI standard deviation (SD) were recorded. Corresponding graded-thickness slices were aligned between systems using saved mean CT numbers and common slice spacing.

For the graded-thickness phantom, four no-attenuator slices were averaged as baseline; subsequent slices were averaged within 1-mm skull-thickness bins. CT-number change was measured relative to baseline; its absolute magnitude was used for the bias analysis. Noise was the within-ROI SD. Absolute contrast was the absolute CT-number difference between two materials. CNR was contrast divided by the root mean square of their within-ROI SDs. All 45 graded-thickness and 28 pediatric material pairs were analyzed. Graded-thickness contrast-gain plots excluded pairs with energy-integrating contrast below 5 HU, leaving 43–45 pairs per bin; CNR retained all pairs. Each dSi-PCCT reconstruction was compared with each energy-integrating scan. The phantoms were analyzed separately.

### Spectral and UHR Assessment

Matched SR and UHR temporal-bone images were selected by closest slice-center location, with a 0.01-mm location difference. Profiles were placed across three high-contrast edges and one trabecular path at corresponding physical coordinates. Each trace was averaged across nine parallel profiles spanning 0.664 mm and smoothed with a 0.12-mm Gaussian window. A four-parameter logistic curve was fitted to each edge profile, and edge width was defined as the distance between the fitted 10% and 90% transition points. Edge profiles were normalized to local plateau levels; the trabecular profile was min-max normalized and was not fitted. SR interpolation to the UHR grid was used for display only; measurements used the native image data.

### Statistical Analysis

Outcomes were summarized by medians and interquartile ranges across ROIs or material pairs after averaging adjacent slices. Mixed-effects models assessed changes with skull thickness and imaging condition. Graded-thickness models used eight nonzero bins for absolute CT-number bias and noise, and baseline plus eight bins for contrast and CNR. ROI or material pair had random intercepts and thickness slopes; pair models accounted for shared ROIs. Pediatric models compared imaging conditions using random intercepts without thickness slopes. Positive outcomes were log transformed. Six dSi-PCCT versus EID comparisons were Holm-adjusted within each outcome. Estimates are reported with 95% confidence intervals and two-sided P values; P < .05 indicated significance. Analyses used MATLAB R2024b (MathWorks).

## RESULTS

### Graded-Thickness Phantom

CNR decreased by 4.17%, 3.25%, and 1.75% per millimeter for the 50-, 65-, and 100-keV dSi-PCCT reconstructions, compared with 8.94% and 8.54% for energy-integrating CT at 80 and 100 kVp (interaction P < .001). Noise increased by 4.54%, 3.07%, and 1.35% per millimeter for dSi-PCCT, compared with 8.38% and 7.84% for energy-integrating CT (interaction P < .001).

Changes in absolute CT-number bias and contrast with skull thickness did not differ significantly among imaging conditions (P = .92 and P = .59, respectively). The 100-keV reconstruction provided the most stable CNR as skull thickness increased, whereas lower-keV reconstructions provided greater absolute contrast (Figure 2).

**Figure 2.**
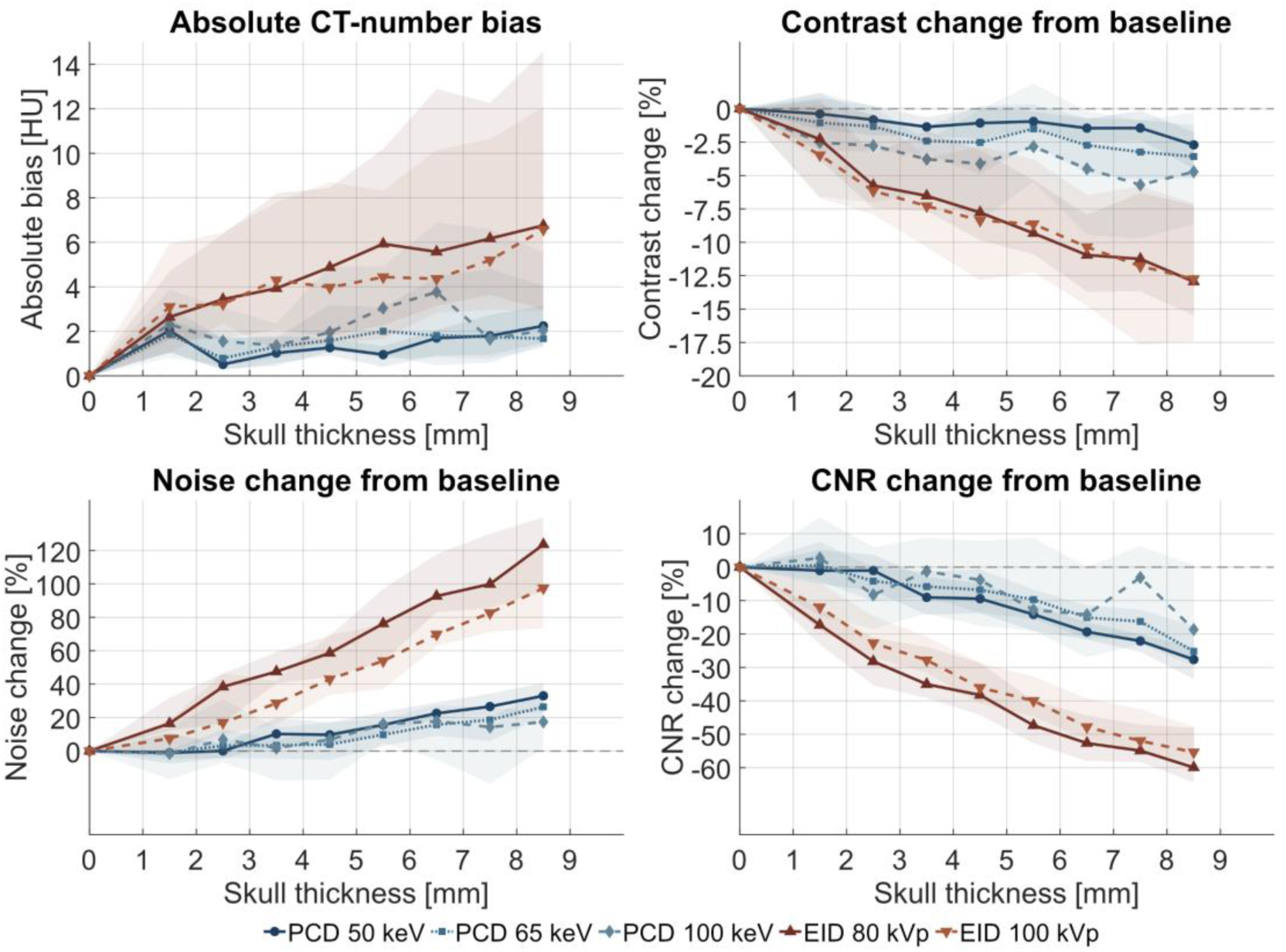
Global quantitative performance across skull thickness in the graded-thickness phantom. Absolute CT-number bias, contrast change from baseline, noise change from baseline, and CNR change from baseline are shown for five imaging conditions. Lines represent medians, and shaded regions represent interquartile ranges across 10 ROIs for CT-number bias and noise and 45 material pairs for contrast and CNR. Zero skull thickness represents the average of four no-attenuator slices; subsequent measurements were averaged within 1-mm bins. Abbreviations: CNR, contrast-to-noise ratio; CT, computed tomography; EID, energy-integrating detector CT; PCD, deep-silicon photon-counting CT; ROI, region of interest.

At 3–4 mm of skull thickness, the median pair-specific contrast ratio at 50 keV was 1.13 relative to 80 kVp and 1.36 relative to 100 kVp. Median pair-specific CNR ratios were 1.79 and 1.71, respectively. The larger CNR advantage was consistent with slower noise growth on dSi-PCCT (Figure 3).

**Figure 3.**
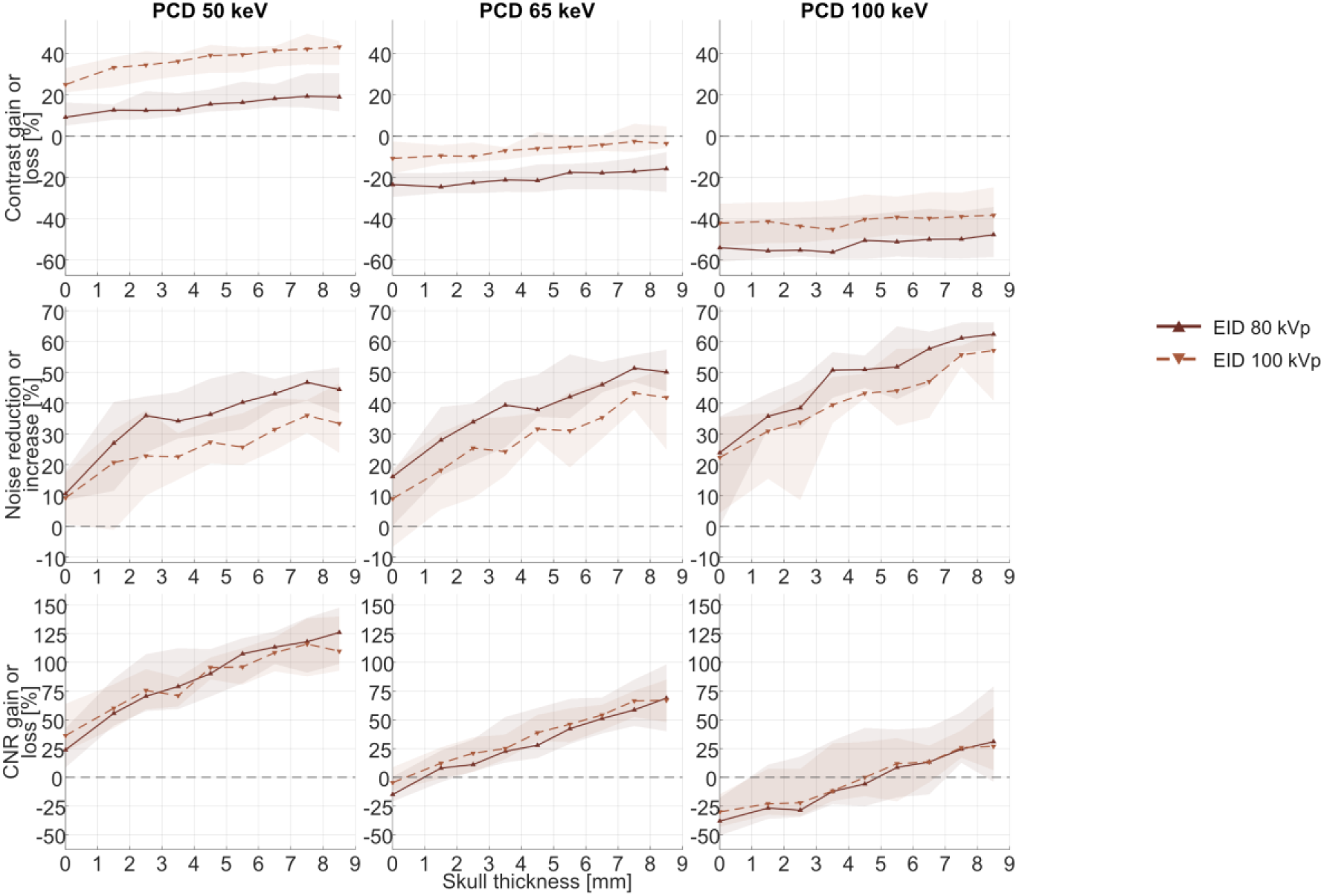
Photon-counting CT gain or loss relative to each energy-integrating acquisition across skull thickness. Columns show the 50-, 65-, and 100-keV PCD reconstructions. Rows show contrast gain or loss, noise reduction or increase, and CNR gain or loss relative to EID-CT at 80 kVp (solid lines) and 100 kVp (dashed lines). Lines represent medians, and shaded regions represent interquartile ranges across 43–45 material pairs for contrast, 45 pairs for CNR, and 10 ROIs for noise. Contrast gains exclude pairs with EID contrast below 5 HU in the corresponding bin. Positive values indicate greater PCD contrast or CNR and lower PCD noise; zero indicates no difference between imaging conditions. Abbreviations: CNR, contrast-to-noise ratio; EID-CT, energy-integrating detector CT; PCD, deep-silicon photon-counting CT; ROI, region of interest.

### Pediatric Head Phantom

The same 50-keV contrast and CNR advantage was observed in the pediatric head phantom. Median pair-specific contrast ratios at 50 keV were 1.09 relative to 80 kVp and 1.33 relative to 100 kVp; median pair-specific CNR ratios were 1.24 and 1.36, respectively (Supplemental Data). Imaging condition was associated with contrast and CNR (both *P* < .001), but not with noise (*P* = .23). Model-estimated 50-keV contrast ratios were 1.097 (95% CI: 1.034, 1.163) relative to 80 kVp and 1.335 (95% CI: 1.259, 1.416) relative to 100 kVp. Corresponding CNR ratios were 1.299 (95% CI: 1.163, 1.452) and 1.424 (95% CI: 1.275, 1.592).

### Spectral and UHR Imaging

One 120-kVp dSi-PCCT acquisition provided SR and UHR 65-keV images and calcium- and water-density maps (Figure 4). The calcium map depicted the mineralized skull, whereas the water map depicted the gel-filled compartment and internal vials.

**Figure 4.**
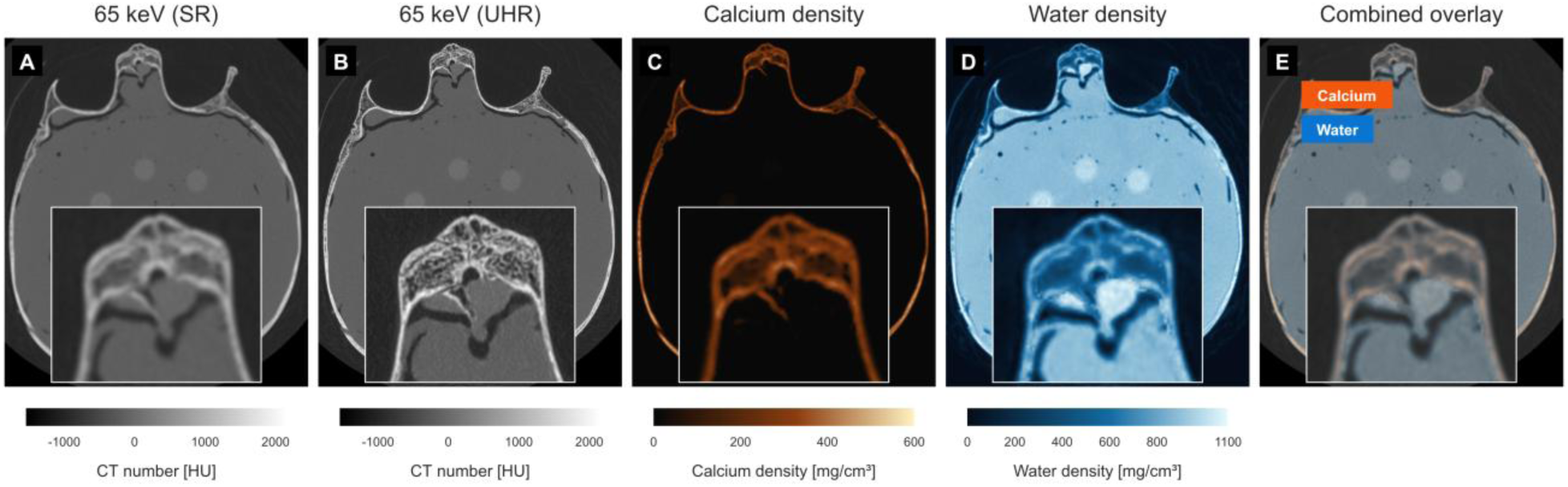
Spectral and ultra-high-resolution imaging of the pediatric head phantom from a single 120-kVp deep-silicon photon-counting CT acquisition. (A) SR and (B) UHR 65-keV images, (C) calcium-density map, (D) water-density map, and (E) combined overlay. Bottom-center insets magnify the same bone region in each panel. Orange and blue denote calcium and water in E. SR, standard resolution; UHR, ultra-high resolution.

At matched axial locations, the UHR reconstruction showed visibly sharper cortical and air-bone interfaces and finer trabecular detail than the SR reconstruction. Fitted 10%–90% transition widths at P1, P2, and P3 were 1.042, 1.223, and 1.791 mm for SR versus 0.431, 0.471, and 0.683 mm for UHR, respectively. UHR widths were 58.6%, 61.5%, and 61.9% narrower (median, 61.5%); fit R² values ranged from 0.978 to 1.000 for SR and from 0.936 to 0.998 for UHR.

Along P4, the UHR trace showed finer local variation through trabecular bone (Figure 5). These band-averaged profiles were descriptive measurements.

**Figure 5.**
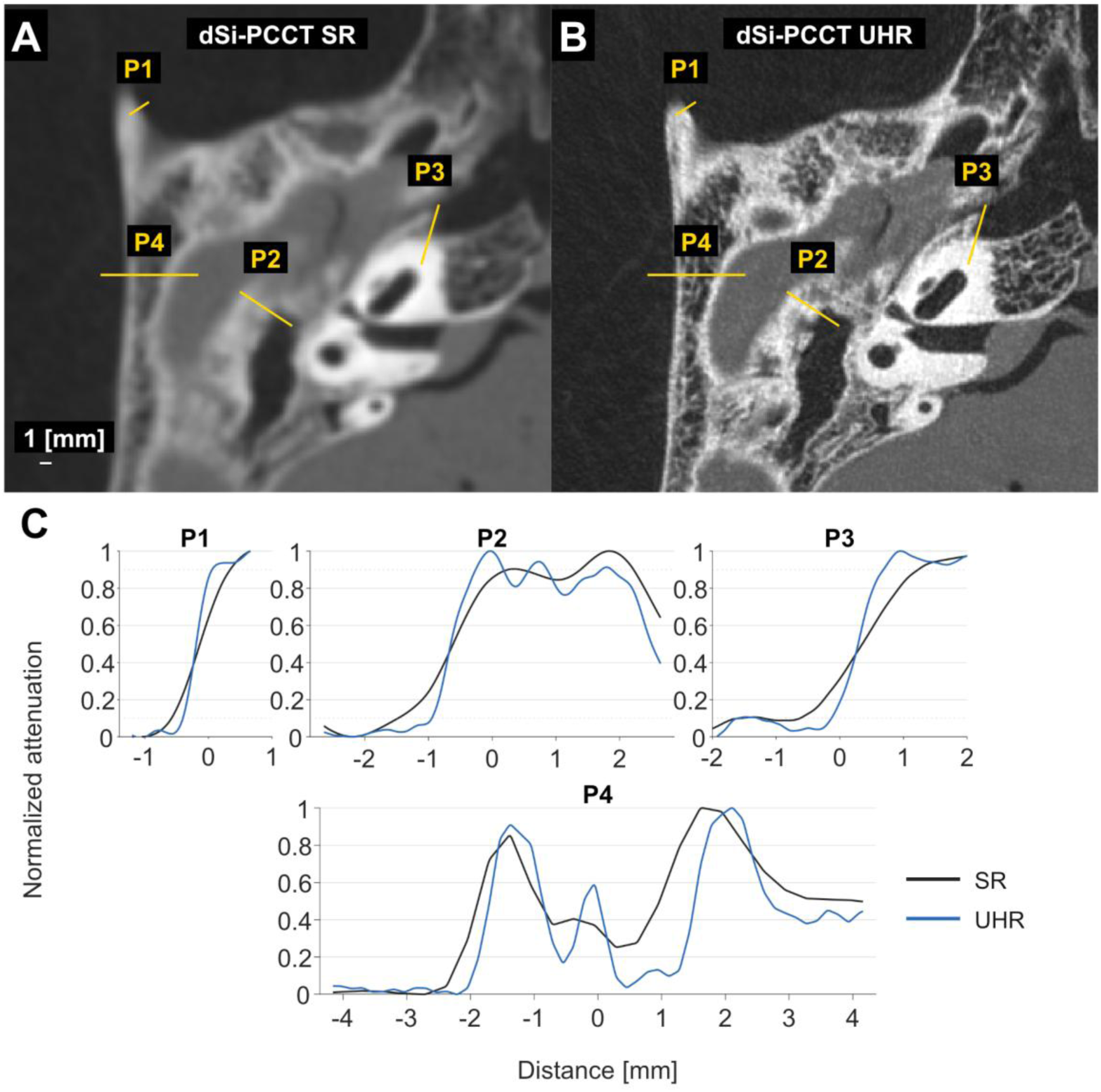
Same-acquisition standard- and ultra-high-resolution temporal-bone imaging. (A) Standard-resolution (SR, HD Bone Kernel) and (B) ultra-high-resolution (UHR; prototype high resolution kernel for UHR mode) 65-keV reconstructions from the same deep-silicon photon-counting CT acquisition. Yellow lines identify profiles P1–P4. Window level/width, 310/3700 HU; scale bar, 1 mm. The SR image was resampled onto the UHR grid for display only; profiles were measured on native images. (C) Matched profiles across three bone edges (P1–P3, top) and a trabecular path (P4, bottom). All plots share the same horizontal distance scale. Black denotes SR and blue denotes UHR. Each curve averages nine parallel traces spanning 0.664 mm. For display, P1 and P3 were normalized to median endpoint levels, with additional scaling to the visible peak for P3 UHR. P2 and P4 were normalized to each curve’s minimum and maximum. Dotted lines mark normalized levels of 0.1 and 0.9.

## DISCUSSION

Deep-silicon photon-counting CT maintained contrast-to-noise ratio more consistently than energy-integrating CT as skull thickness increased, primarily because noise increased more slowly. Changes in contrast with increasing skull thickness did not differ significantly among imaging conditions. The complementary spectral outputs served different purposes: 50 keV provided the greatest absolute contrast and contrast-to-noise ratio, whereas 100 keV provided the most stable contrast-to-noise ratio across increasing attenuation. Because all three reconstructions arose from one scan, image contrast could be selected after acquisition for the diagnostic task ^2,3^.

The pediatric head phantom showed the same reconstruction-dependent contrast–noise pattern, including a 50-keV CNR advantage over both energy-integrating conditions. Exact ratios were not expected to match because the phantoms differed in geometry, vial composition, and vial position. Concordance across the controlled and anatomically realistic experiments supports further study of one dSi-PCCT acquisition across pediatric head sizes and tasks ^14^.

The matched temporal-bone comparison demonstrated that the same dSi-PCCT acquisition supported both spectral and 65-keV UHR reconstruction. UHR showed sharper selected edge transitions and finer trabecular detail than SR, consistent with prior temporal-bone photon-counting CT studies ^8,9^. The profiles demonstrate local edge sharpness within the selected anatomy.

We assessed attenuation-dependent behavior without activating optional bone-specific or additional beam-hardening correction. Such correction could reduce CT-number deviations on either system. Prior simulation and phantom studies found different and, in some settings, greater beam-hardening effects with photon-counting than energy-integrating detection, while spectral reconstruction permitted post-acquisition mitigation ^15,16^. Correction performance across pediatric head sizes was not evaluated.

This study had limitations. It compared one scanner of each detector type without repeated scans, and detector, acquisition, and reconstruction effects could not be separated. The concrete attenuator did not reproduce the composition or geometry of pediatric bone. Vial materials were heuristic attenuation references, and neither phantom represented the full range of patient anatomy. Representative image pairs and anatomical edge profiles do not establish diagnostic performance or fully characterize spatial resolution.

In conclusion, deep-silicon photon-counting CT maintained contrast-to-noise ratio more consistently than energy-integrating CT as skull thickness increased. One acquisition provided complementary spectral and ultra-high-resolution outputs, supporting further evaluation for quantitative and high-resolution pediatric head imaging.

## DECLARATIONS

### Ethics approval

Not applicable. This phantom study involved no living participants or identifiable human data.

## Previous presentation

A related neonatal brain custom-phantom abstract was presented at the 2026 Joint AAPM | COMP Annual Meeting & Exhibition, Vancouver, BC, Canada, July 19, 2026 (SU-315-GPD-D-180).

## Data Availability

Data generated or analyzed during this study are available from the corresponding author upon reasonable request.

## ACKNOWLEDGMENTS

We used OpenAI Codex (GPT-5.6 and GPT-6; OpenAI) to assist with manuscript drafting and editing, MATLAB code development, figure and table preparation, and citation organization and formatting. The corresponding author reviewed and verified all AI-assisted content and outputs. The authors take full responsibility for the accuracy and integrity of the work.

## Abbreviations

CNR: contrast-to-noise ratio
CTDIvol: volume CT dose index
DL-M: medium-strength deep-learning reconstruction
dSi-PCCT: deep-silicon photon-counting CT
EID: energy-integrating detector
EID-CT: energy-integrating detector CT
HU: Hounsfield unit
MTF50/MTF10: spatial frequencies at 50%/10% of the modulation transfer function
PCD: deep-silicon photon-counting CT
SD: standard deviation
SR: standard resolution
UHR: ultra-high resolution

## REFERENCES

1. Cauley KA, Yorks PJ, Flora S, Fielden SW. The effects of the skull on CT imaging of the brain: a skull and brain phantom study. Br J Radiol. Mar 1 2021;94(1119):20200714. doi:10.1259/bjr.20200714

2. Michael AE, Boriesosdick J, Schoenbeck D, et al. Image-Quality Assessment of Polyenergetic and Virtual Monoenergetic Reconstructions of Unenhanced CT Scans of the Head: Initial Experiences with the First Photon-Counting CT Approved for Clinical Use. Diagnostics (Basel). Jan 21 2022;12(2):265. doi:10.3390/diagnostics12020265

3. Michael AE, Schoenbeck D, Woeltjen MM, et al. Nonenhanced Photon Counting CT of the Head: Impact of the keV Level, Iterative Reconstruction and Calvaria on Image Quality in Monoenergetic Images. Clin Neuroradiol. Mar 2024;34(1):75–83. doi:10.1007/s00062-023-01331-w

4. Michael AE, Petersilka M, Schoenbeck D, et al. Non-contrast photon counting computed tomography of the head: optimized modeling of off-focal radiation to reduce calvarium-related tissue inhomogeneity. BMC Med Imaging. Jul 18 2025;25(1):289. doi:10.1186/s12880-025-01718-w

5. Smith K, Politte D, Reiker G, et al. Automated measurement of pediatric cranial bone thickness and density from clinical computed tomography. Annu Int Conf IEEE Eng Med Biol Soc. 2012;2012:4462–5. doi:10.1109/embc.2012.6346957

6. Willemink MJ, Persson M, Pourmorteza A, Pelc NJ, Fleischmann D. Photon-counting CT: technical principles and clinical prospects. Radiology. 2018;289(2):293–312.

7. Pourmorteza A, Symons R, Reich DS, et al. Photon-Counting CT of the Brain: In Vivo Human Results and Image-Quality Assessment. AJNR Am J Neuroradiol. Dec 2017;38(12):2257–2263. doi:10.3174/ajnr.A5402

8. Zhou W, Lane JI, Carlson ML, et al. Comparison of a Photon-Counting-Detector CT with an Energy-Integrating-Detector CT for Temporal Bone Imaging: A Cadaveric Study. AJNR Am J Neuroradiol. Sep 2018;39(9):1733–1738. doi:10.3174/ajnr.A5768

9. Lee JS, Kim J, Bapuraj JR, Srinivasan A. Comparison of Image Quality and Radiation Dose in Pediatric Temporal Bone CT Using Photon-Counting Detector CT and Energy-Integrating Detector CT. AJNR Am J Neuroradiol. Sep 9 2024;45(9):1322–1326. doi:10.3174/ajnr.A8276

10. Grunz JP, Heidenreich JF, Lennartz S, et al. Spectral Shaping Via Tin Prefiltration in Ultra-High-Resolution Photon-Counting and Energy-Integrating Detector CT of the Temporal Bone. Invest Radiol. Dec 1 2022;57(12):819–825. doi:10.1097/rli.0000000000000901

11. Chang S, Benson JC, Lane JI, et al. Ultra-High-Resolution Photon-Counting-Detector CT with a Dedicated Denoising Convolutional Neural Network for Enhanced Temporal Bone Imaging. AJNR Am J Neuroradiol. Jun 3 2025;46(6):1188–1195. doi:10.3174/ajnr.A8572

12. Danielsson M, Persson M, Sjölin M. Photon-counting x-ray detectors for CT. Phys Med Biol. Jan 29 2021;66(3):03TR01. doi:10.1088/1361-6560/abc5a5

13. Fredenberg E, Collin D, Carbonne L, Wu M, Man B, Gronberg F. Simulating and correcting the pileup effect in deep-silicon photon-counting CT. Med Phys. Aug 2025;52(8):e18075. doi:10.1002/mp.18075

14. Kluner LV, Zensen S, Peuster H, et al. Radiation dose and signal-to-noise ratio in pediatric head CT: a phantom study comparing photon-counting and energy-integrating detectors. Neuroradiology. Jul 2026;68(7):1811–1823. doi:10.1007/s00234-026-04075-9

15. Shikhaliev PM. Beam hardening artefacts in computed tomography with photon counting, charge integrating and energy weighting detectors: a simulation study. Phys Med Biol. Dec 21 2005;50(24):5813–27. doi:10.1088/0031-9155/50/24/004

16. Pourmorteza A, Choux AR, Holmes TW, et al. Beam hardening of K-edge contrast agents: a phantom study comparing clinical energy-integrating detector and photon-counting detector CT systems. Eur Radiol Exp. Mar 19 2025;9(1):31. doi:10.1186/s41747-024-00530-5

